# Do community pharmacies add value to immunization programs? A systematic review from the UK between 2015-2025

**DOI:** 10.1101/2025.09.12.25335325

**Authors:** KA Thomas, SA Ismail, T Chantler, S Perman, J Pryce

## Abstract

**Background:** Community pharmacies (CPs) have played an increasing role in the delivery of influenza and COVID-19 vaccination in recent years. This systematic review updates an analysis conducted in 2015, looking at the acceptability and cost-effectiveness of CPs and their impact on inequalities and improving vaccine uptake.

**Methods:** We searched MEDLINE, Embase, Scopus, Web of Science and CINAHL (09/10/2025) for English-language papers (2015–25) and identified unpublished literature through NHS regional teams, UKHSA, and pharmacy associations. Studies on NHS-commissioned community pharmacy immunization services were included, private/travel vaccinations excluded. Quality of identified literature was appraised using the Mixed Methods Appraisal Tool. The AACODS checklist was completed for non-peer-reviewed grey literature. A narrative synthesis categorized outcomes into acceptability, vaccine uptake, cost-effectiveness, and impact on inequalities.

**Results:** Eleven peer-reviewed articles and five grey literature reports of evaluations of CP delivered influenza, COVID-19, Measles-Mumps-Rubella, Mpox, Polio and Respiratory Syncitial Virus (RSV) immunisation programmes were included. While CPs account for a growing share of vaccination delivery for certain seasonal programmes, there was no evidence of an increase in coverage rates or reduced inequity in vaccine uptake. User acceptability of CP-delivery was high, with convenience and accessibility cited. The few studies reporting comparator data suggest similar levels of favourability to GP-led delivery. Barriers included fragmented data systems, logistical constraints, and high wastage rates. No studies included evidence on cost-effectiveness. Some papers suggested CPs had the potential to reach underserved and multi-ethnic communities, but there was a lack of robust evidence for this.

**Conclusions:** CPs can support immunisation delivery, with impact varying by context. There is no cost-effectiveness analysis that incorporates a comprehensive assessment of all relevant clinical, economic, and contextual factors. Evidence suggests CPs may strengthen capacity during periods of peak demand but require better integration and support to improve equity and contribute meaningfully to routine vaccination programme delivery.

**Key messages:** *What is already known on this topic:* Community pharmacies (CPs) have increasingly contributed to vaccination delivery in the UK, particularly for influenza and COVID-19, but the broader impact, cost-effectiveness, and equity implications of CP-led programmes remain unclear. An earlier review published in 2015 highlighted limited evidence on these outcomes.

*What this study adds:* This systematic review finds that while CPs play a growing role in vaccination delivery, there is no robust evidence that this has improved overall vaccine uptake or reduced health inequalities. Acceptability among users is high, but reported issues include poor data integration and logistical barriers. The lack of cost effectiveness evaluations limit assessment of impact and scalability.

*How this study might affect research, practice or policy:* Findings highlight the need for more comprehensive evaluations of CP-led vaccination services, particularly on cost-effectiveness and equity impact. Policymakers should consider investing in better integration and infrastructure to support CPs in contributing more meaningfully to routine and targeted immunisation efforts.

## Introduction

Vaccination programmes are highly effective public health interventions that prevent infectious disease, reduce health inequalities and have been shown to have averted millions of deaths worldwide (1). In the United Kingdom (UK), routine immunisations are typically administered in general practice (GP) and educational settings (for school-age immunisations). Following the introduction of the National Health Service (NHS)-commissioned Community Pharmacy (CP) Flu vaccination service in England in 2015, some immunisations are also administered in CPs.

CPs are an integral part of the UK healthcare system, providing a wide range of public services (2). Traditionally, they are responsible for medication dispensing, patient advice and health promotion activities, and Pharmacy First services (which cover care provision for defined, minor conditions, and provision of urgent repeat medications) (3). More recently, CPs have been integrated into delivery arrangements for seasonal vaccinations (influenza and COVID-19) in the UK (4).

Arguments presented in support of CP-based vaccination delivery have emphasised perceived convenience, and potential benefits for working-age adults and underserved populations (5, 6). Features of the CP model that have been identified as advantageous include pharmacists’ position as recognised healthcare providers, the potential to educate patients about vaccines (7), and perceived ability to reach individuals who might not get key vaccinations due to limited time or healthcare access barriers (8). CPs can be found on most high streets and in many rural communities and often offer extended opening hours (9). They played an important role during the COVID-19 pandemic, where they were integrated into the national delivery strategy for vaccinations. The unprecedented expansion of CP vaccination services globally during the COVID-19 pandemic demonstrated that pharmacies could function as high-volume immunization providers, although the circumstances and resourcing available to support this were unique and may not be applicable to routine immunisation programmes (10).

However, the overall impact of CPs on vaccination delivery is uncertain, largely due to lack of evidence and inconsistent findings across studies. A previous review of the acceptability and impact of CP immunisation programmes in the UK was published in 2018, including data from peer-reviewed studies published before November 2015. The review identified three peer-reviewed studies and 25 unpublished reports, each of which described seasonal influenza programmes. Overall, the review found no evidence that CPs increased vaccination uptake, and limited data to suggest they improved access for individuals who had not been vaccinated before (6). That analysis found no evidence on cost-effectiveness of delivery via CPs.

Given the expansion in both scale and scope of CP vaccination services over the last decade, this review aimed to build on these findings by evaluating the evidence of all CP-led routine immunization programmes in the UK between 2015 and 2024 (6). We sought evidence on effects on vaccine uptake, public and provider acceptability, cost-effectiveness, and equity in vaccine access. Findings were intended to inform policy considerations as to whether CPs should take on a broader, long-term role in national immunization strategies.

## Methods

We followed the Preferred Reporting Items for Systematic Reviews and Meta-Analyses (PRISMA) guidelines (11) and a review protocol registered with the PROSPERO international database of prospectively registered systematic reviews in health and social care (CRD42025622975). We conducted keyword-structured searches on MEDLINE, Embase, Scopus, Web of Science and CINAHL on 9th October 2025 for peer-reviewed papers published in the English language since November 2015. Details of the search strategy can be found in Supplementary File 1. We also identified unpublished literature by contacting the 1) screening and immunization leads in each of the NHS regional teams in England; 2) the National Immunisation Programmes Division at UKHSA; and 3) The Company Chemists’ Association (CCA), a trade association representing CPs in the UK, to ask for evaluations of local immunisation programmes. The cut-off date for responding was 17^th^ January 2025. Documents that reported on locally or nationally NHS commissioned CP services which offered free vaccines as part of the national immunization schedule, and as part of the pandemic, were included in the review. Papers focused solely on travel vaccinations or private vaccinations were excluded.

A two-stage screening process was conducted using Rayyan AI software. Titles and abstracts were screened to remove non-pertinent studies prior to conducting full text assessments of all potentially relevant studies. Each screening stage was conducted by two reviewers independently (KT and JP), with any discrepancies resolved through discussion. Data extraction was performed by one of two reviewers (KT or JP) using a predefined template in Microsoft Word which included the following variables: publication type, setting, year, study design, vaccine type, target group, scale of implementation, and reported outcomes. Missing data were requested from study authors via email.

The quality of each identified study was assessed using the Mixed Methods Appraisal Tool (MMAT), which classifies papers into qualitative research, randomized controlled trials, non-randomized studies, quantitative descriptive studies, or mixed-methods studies to apply relevant criteria for assessment (12). Unpublished grey literature articles that did not meet any of the above categories, such as case studies or industry reports, were critically appraised using the AACODS checklist (13).

Due to anticipated heterogeneity in programme objectives, implementation methods, and evaluation strategies, a narrative synthesis was performed. Synthesized data were structured around the review’s four evaluation domains: acceptability, vaccine uptake, cost-effectiveness, and impact on inequalities. Narrative synthesis focused on describing the variety of outcomes that were reported for each domain, the extent to which results of studies were consistent with one another, whether any heterogeneity was influenced by characteristics of different study design and evaluation methodologies, and the certainty of the evidence provided.

## Results

### Results of the search

The PRISMA flow diagram in **Figure 1** describes the results of the search. In total, 1,314 articles were identified through database searches. After removal of duplicates, 810 abstracts were screened for eligibility, and 21 articles were identified as potentially relevant. After assessing the full text articles against the review inclusion criteria, we identified 11 peer-reviewed studies (2, 4, 14–22) and 11 grey literature articles relating to five different CP vaccination service evaluations (23–27) (**see table 1 and table 2**).

**Figure 1:**
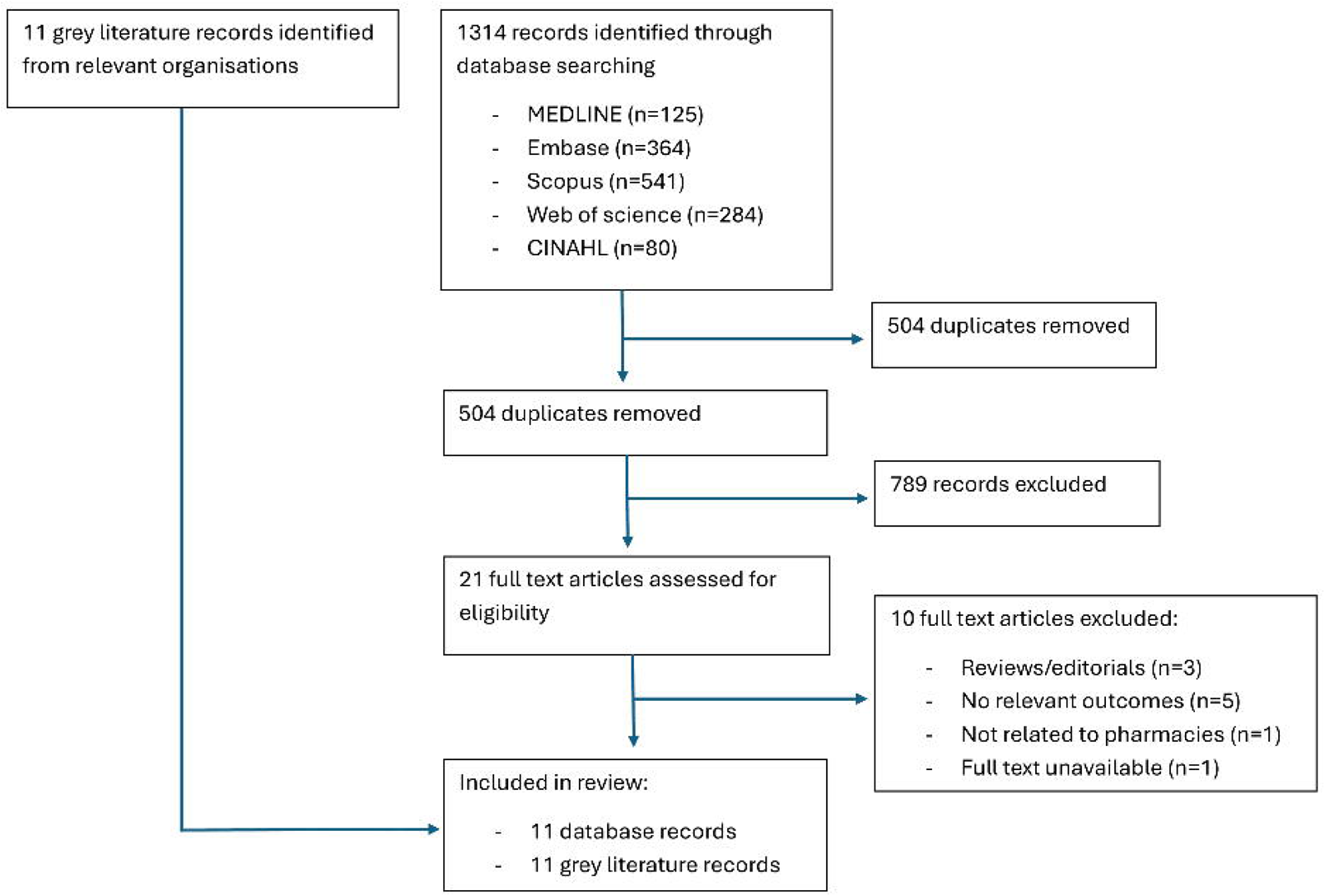

**Table 1:** Key features of included published literature and grey literature.

|  |  |
| --- | --- |
| 11 published papers and 5 grey literature | N (%) |
| **Methodological approach** |  |
| Quantitative | 11 (69%) |
| Qualitative | 3 (19%) |
| Mixed Methods | 2 (13%) |
| **Vaccine Type** |  |
| Influenza | 7 (44%) |
| Covid-19 | 6 (38%) |
| MMR | 1 (6%) |
| Mpox | 1 (6%) |
| Polio | 1 (6%) |
| RSV | 1 (6%) |
| **Target Group** |  |
| All eligible ages and risk groups for free flu vaccine | 5 (31%) |
| All eligible ages and risk groups for COVID-19 vaccination | 4 (25%) |
| All those aged 5+ eligible for MMR vaccination | 1 (6%) |
| 5-6 year-olds | 1 (6%) |
| 1–9 year-olds | 1 (6%) |
| 18–29 year-olds | 1 (6%) |
| 75–79 year-olds | 1 (6%) |
| Pregnant women | 1 (6%) |
| Gay, bisexual and other men who have sex with men | 1 (6%) |
| **Setting** |  |
| London | 3 (19%) |
| Wales | 2 (13%) |
| West Midlands | 1 (6%) |
| Multiple regions in England | 3 (19%) |
| United Kingdom | 2 (13%) |
| Southwest England | 1 (6%) |
| Northwest England | 1 (6%) |
| East of England | 2 (13%) |
| Northern Ireland | 1 (6%) |
| **Outcomes reported** |  |
| Uptake | 11 (69%) |
| Acceptability | 12 (75%) |
| Cost | 6 (38%) |
| Inequalities | 8 (50%) |

**Table 2:**
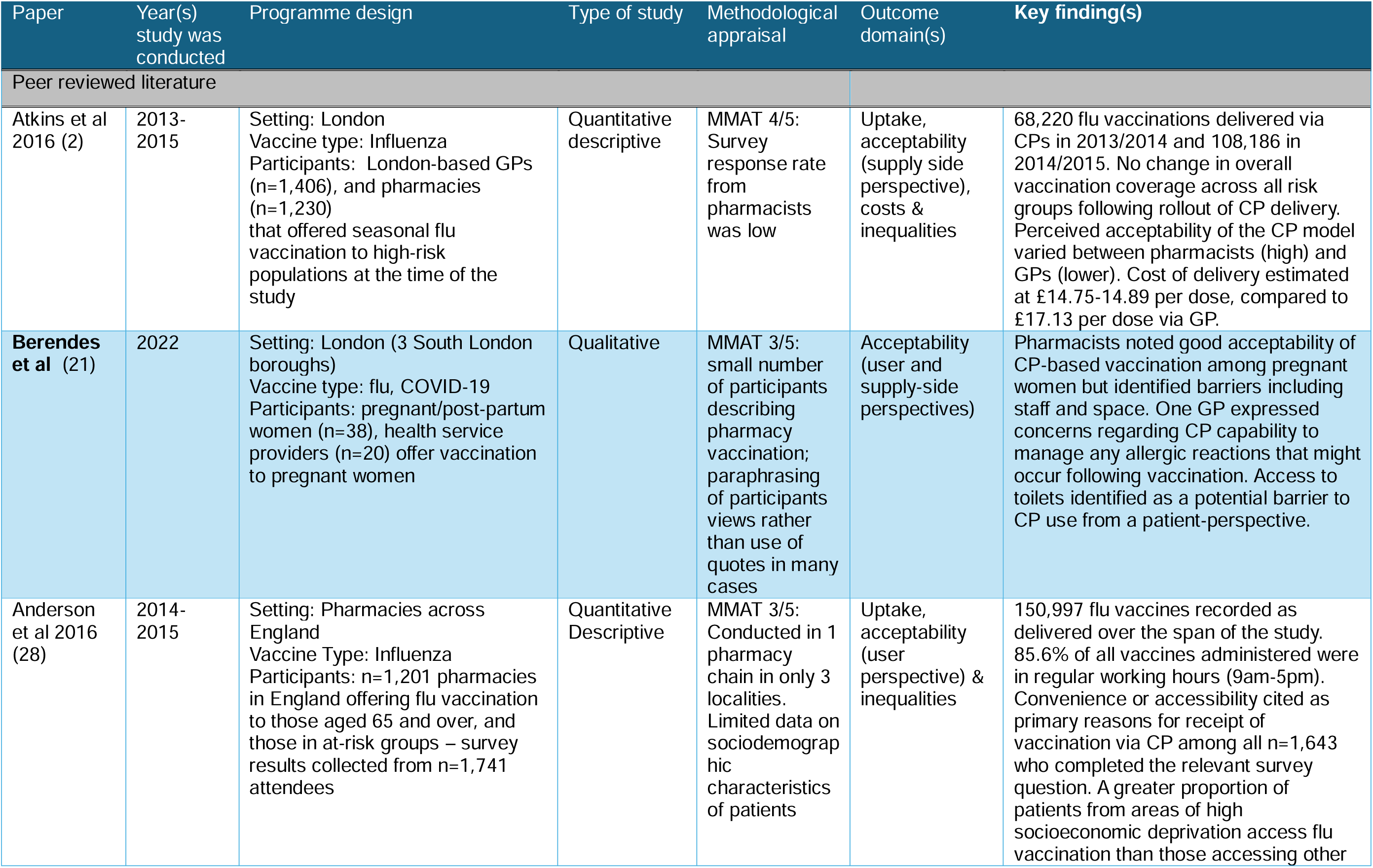
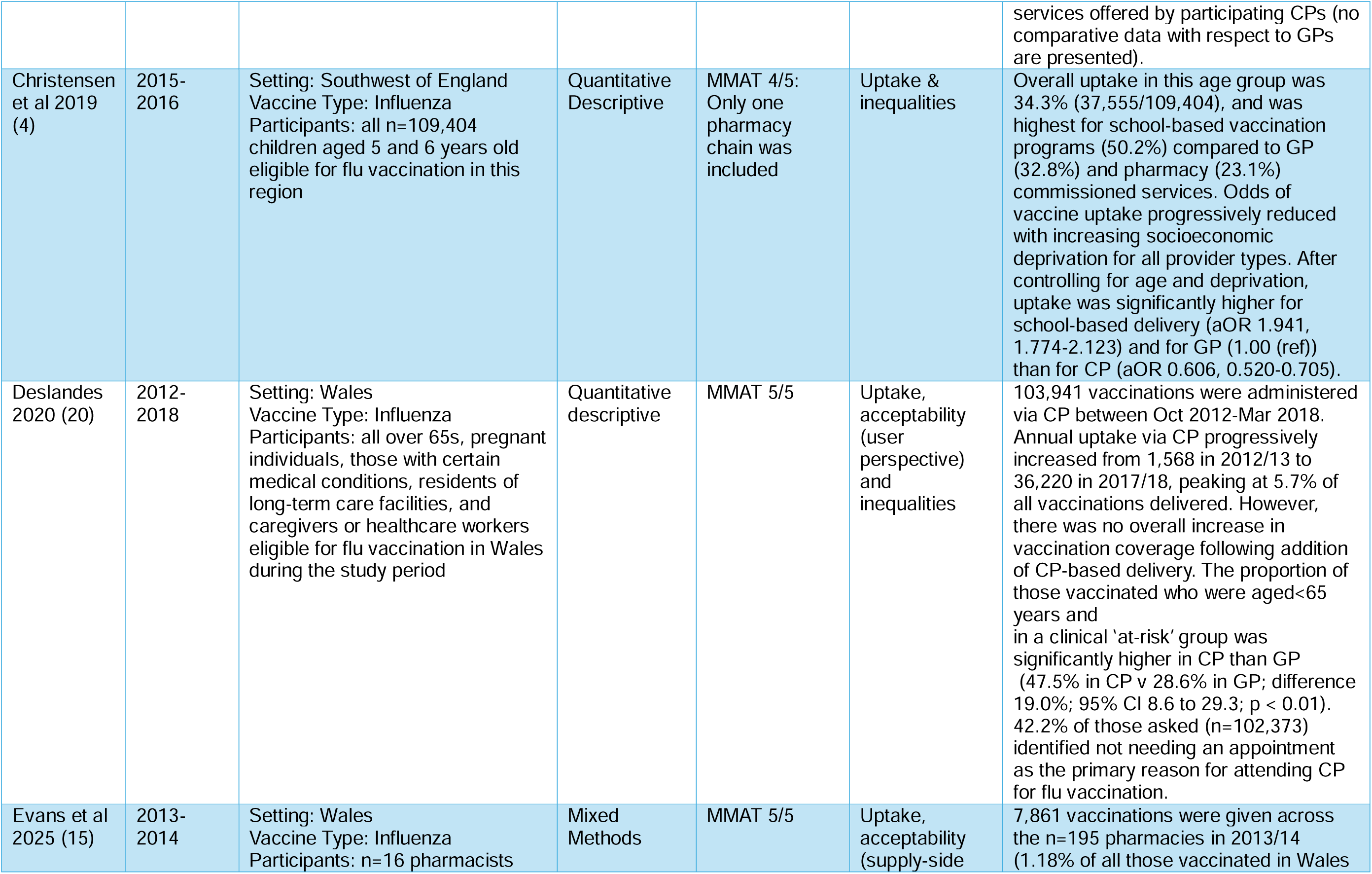
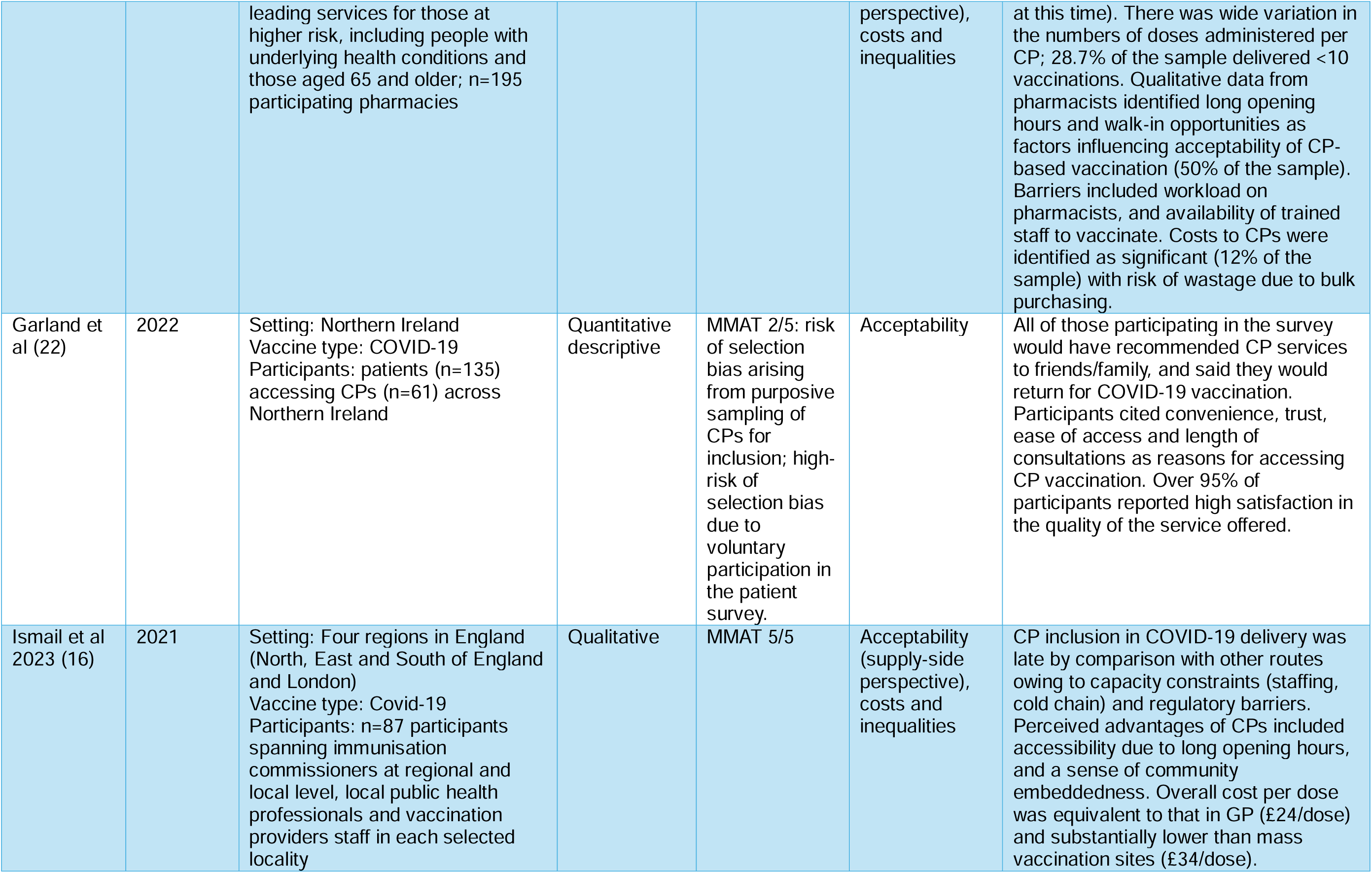
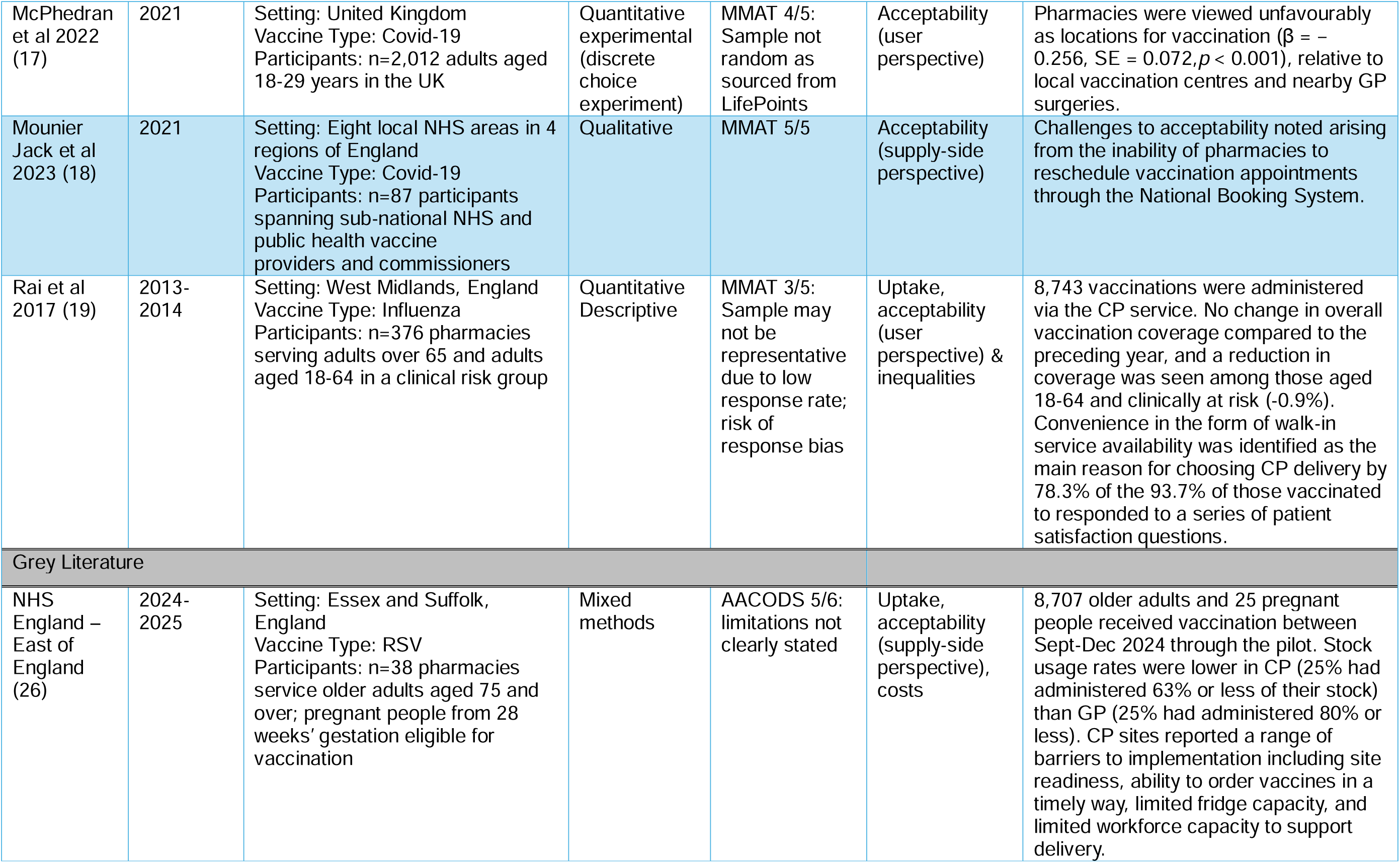
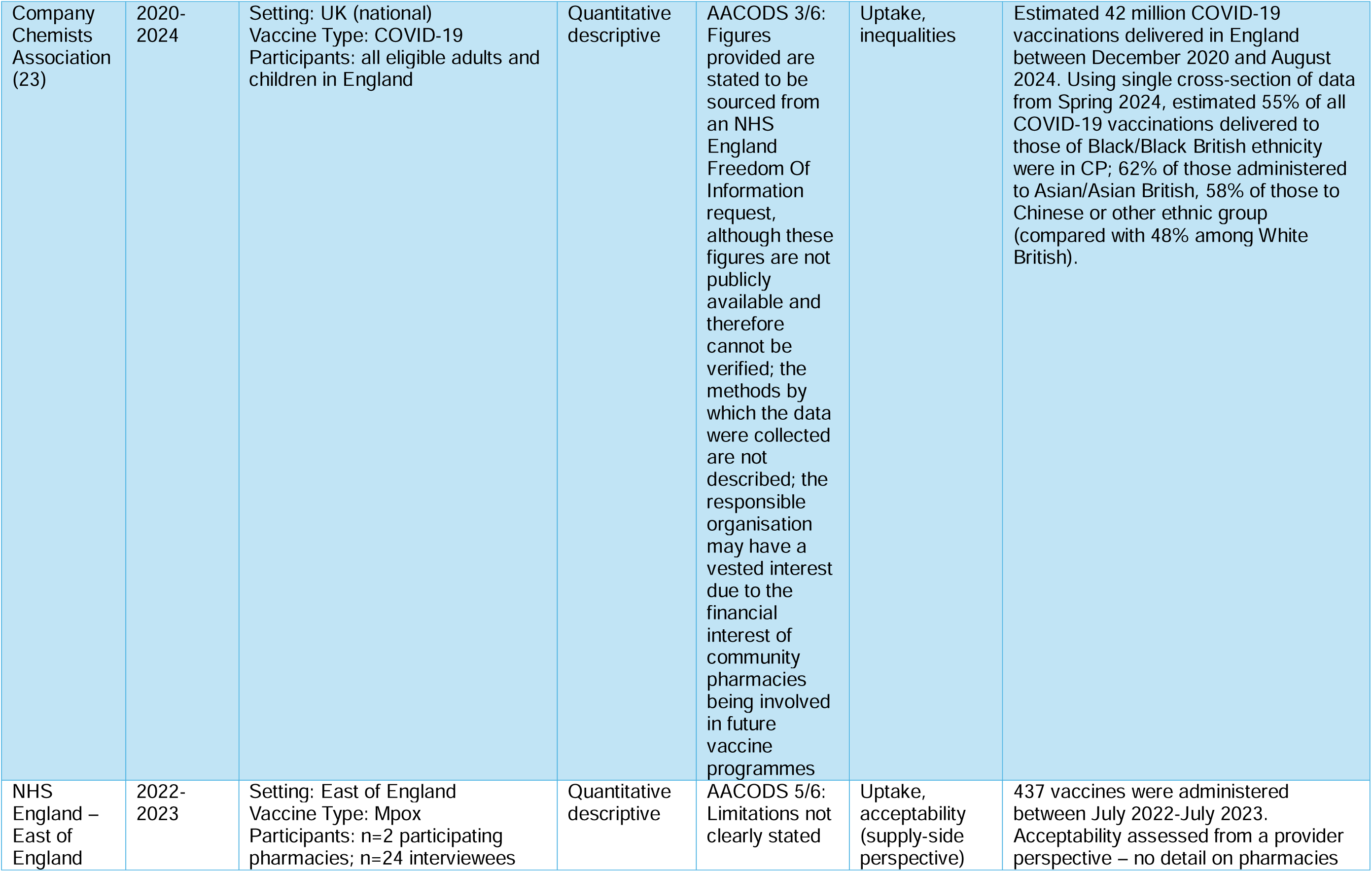
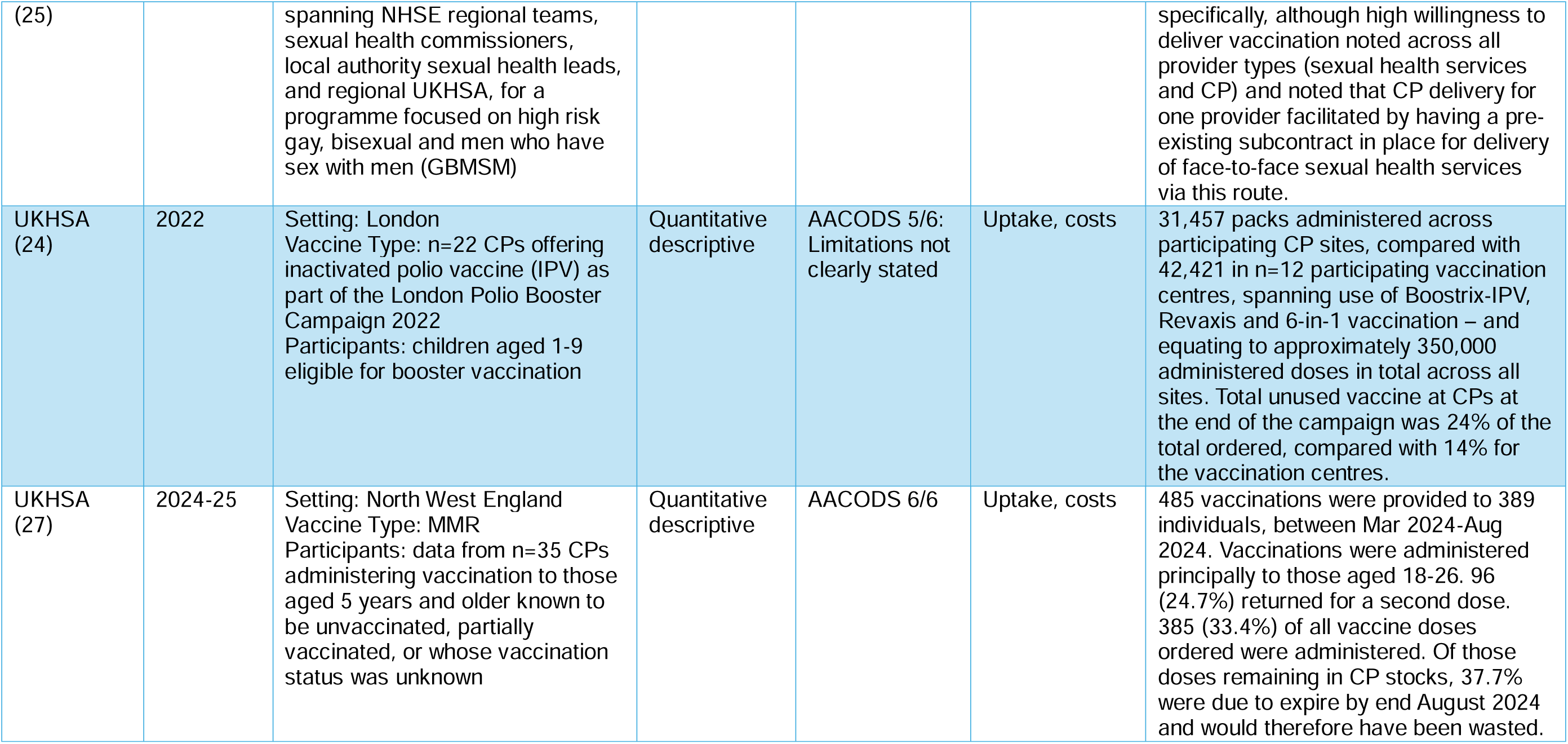
Summary of included published literature.

### Characteristics and quality of included studies

The characteristics of the included studies are described in **table 1**. Of the 11 peer-reviewed articles, six evaluated CP influenza vaccination programmes (2, 4, 15, 19, 20, 28) and four papers related to COVID-19 vaccination services (16–18, 22). The final article included information on the provision of maternal vaccinations in CP, which at the time of the study would have included influenza and COVID-19 (21). Three of the sources considered administration to age groups eligible for routine immunisation (4, 24, 27). The included studies provided data from national programmes in England (17, 28) and Wales (15, 20), and regional programmes in London (2, 21), the West Midlands (19), Southwest England (4), and Northern Ireland (22).

The grey literature included documentation on the national COVID-19 immunisation programme, and regional immunisation programmes in the north-west (Measles-Mumps-Rubella; MMR), London (polio), and the south-east (Mpox and Respiratory Syncytial Virus, RSV) (23–27). The format and quality of the grey literature varied considerably. A service review report of the RSV CP vaccination programme in the East of England contained vaccination supply data and qualitative evaluation data. For CP polio vaccine catch-up in London, only supply data, with the number of vaccinations delivered and administered, were available. For practical purposes, all sources pertaining to individual evaluations were treated as single outputs. The grey literature provided more details on the logistics and implementation of CP vaccination delivery than published papers.

The quality of peer reviewed articles varied. Sampling bias occurred in six articles, due to limitations of sampling approach or low survey response rates (2, 4, 17, 19, 22, 28). Two articles only included data from one pharmacy chain which limited the representativeness of included data (4, 28).

### Uptake

Six out of 9 of the published articles and all five grey literature documents reported data on uptake (2, 4, 15, 19, 20, 23–28). Predominantly these were reported as raw uptake figures without comparators. Most peer-reviewed papers in this group showed increases in the absolute volumes of vaccination administered via CPs over time, although where reported final totals represented a small share of overall vaccination delivery.

The majority of available data on uptake by volume was for the influenza vaccine, and volumes varied considerably by study. For example, in Wales, 7,861 influenza vaccinations were administered across 195 pharmacies (27.3% of all pharmacies), representing 1.18% of total vaccinations in 2013-14 (20). Between 2012-18, a total of 103,941 influenza vaccines were administered in Wales in CPs, with the proportion of vaccines administered in CPs increasing from 0.3% to 5.7% of all vaccinations given (20). In England, 1,201 pharmacies (of around 12,000 in operation at that time (29)) administered a total of 150,997 influenza vaccines during the 2014/2015 season (28). An industry report estimated that CPs delivered 42 million COVID-19 vaccinations between December 2020 and August 2024, accounting for over a quarter of the vaccines administered during this period (23). Critical appraisal of this reference using the AACODS checklist highlighted several risks, particularly as the sources cited in support of these figures are not publicly available and the methods by which they were collected are not described. In the East of England, 8,707 older adults and 25 pregnant individuals received the RSV vaccine via a CP pathfinder from September-December 2024, of over 170,000 doses delivered in this region via all routes during this period (26, 30).

Of the four studies that reported effects of CP delivery on overall coverage, none showed any effect (2, 4, 19, 20). In London, overall influenza vaccine coverage remained similar at around 60% between 2011 to 2014 (2). Coverage varied considerably between high-risk groups, at 70% for over 65s and only 35% for pregnant women; for comparison, national figures for these groups for England in 2013/14 were around 73% and 40% respectively (31). An analysis of influenza vaccination uptake in Wales showed no effect on coverage after introduction of CP-based delivery (20). A study in the West Midlands recorded a total of 8,743 influenza vaccinations administered via CPs in 2014/2015, but uptake did not increase from 2013 (prior to CP commissioning) to 2015 (19). A comparative analysis of school, GP or CP delivery of influenza vaccines for 5-6 year olds during the 2015-2026 season reported that school-based programmes achieved the highest uptake (50.2%), followed by GPs (32.8%) and CPs (23.1%) (4).

Grey literature sources described how CPs have been deployed to support targeted vaccination campaigns in England – but these sources report raw uptake figures without comparators, and in one case use of vaccination packs rather than doses administered. In a northwest England CP pilot MMR catch up campaign, 485 MMR doses were given across 35 participating CPs between March and August 2024 (27). London’s polio campaign in 2022-23 administered 31,457 vaccine packs across 22 CP sites whereas 42,421 were delivered across 12 participating vaccine centres (24).

### Acceptability

Most included literature reported on the acceptability of CPs as a delivery model from the perspective of service users (17, 19, 20, 22, 28) and supply-side actors (2, 15, 16, 18, 25, 26). Where user data were reported, convenience (in particular, the ability to walk-in for vaccination rather than needing an appointment) and accessibility were cited as the main reasons for choosing to get vaccinated at CP by attendees (17, 19, 20, 28), although other factors including trust were also cited (22). In the only study that reported relevant primary, comparative data, 95.8% of patients rated vaccine administration via CP to be as good as administration by a GP or nurse – suggesting comparably high acceptability ratings for CP to other primary care routes (19). However, a discrete choice experiment to test preferences for different, recognised and hypothetical service delivery routes, and involving just over 2,000 adults aged 18-29 found that CPs were viewed significantly less favourably as access points for vaccination than vaccination centres and GP surgeries (17). Comparative acceptability relative to other service points was not reported in the other literature.

Supply-side perspectives were variable and contingent on the source consulted. Analysis of a pilot of seasonal flu vaccination via CPs in England found that participating pharmacies viewed CP-based flu vaccination delivery more favourably than GPs; 98% of CPs and 60% of GPs believed it improved patient choice, and 97% and 40%, respectively cited increased convenience (2). Findings from interviews with pharmacists reported extended hours, urban locations, and walk-in availability as strengths of the CP model (15). However, the only study to report on timing of administration noted that the vast majority of doses were given within regular working hours i.e. 9am-5pm (28). Links with local communities and the presence of staff in CPs with a mix of language skills were noted as helpful for reaching diverse populations by service providers in an evaluation of COVID-19 vaccination delivery models (16). Included literature noted several challenges to CP-based delivery. Research in London and the West Midlands reported that the multiple systems for recording flu vaccination data were not only time consuming to complete but led to a loss of data (2, 19). In addition, workload, availability of trained staff, appropriate space for vaccine administration, costs, and other logistical challenges, such as cold chain storage, were identified as barriers to CP delivery (16, 21, 25, 26). Resistance from local GPs was widely reported with gaps in collaboration portrayed as hindering CP performance. GP concerns regarding the CP delivery model included data leakage, safety and financial losses (2).

### Cost

Seven studies across both the grey and published literature included data on costs, though none reported on cost-effectiveness (2, 15, 16, 24, 26, 27). The cost profile for CP vaccine programmes varied across studies. One study found that while COVID-19 vaccination costs per dose were lower at CPs (£24) than at mass vaccination sites (£34), CP and GP costs per dose were comparable (16). For flu vaccinations in London, CP-administered vaccines were reported to be up to £2.35 less per dose than GP-administered ones (2). However, the CP cost estimates only included reimbursement costs and did not factor in wider costs associated with vaccine programme delivery including the administration of booking systems, and call-recall for registered patients which GPs are expected to deliver as part of the GP contract in England but are frequently provided nationally for CPs. Vaccine wastage was identified as a challenge in a mixed methods study of influenza vaccination delivery in Wales, in which pharmacies reported that wastage contributed to financial losses (15).

Grey literature reported on costs indirectly, by presenting data on high rates of vaccine wastage. This was identified as a challenge to CP delivery by three of the five included service evaluations. In an MMR pilot in northwest England, only 39.6% of ordered MMR vaccines were used, and 33.7% of doses ordered by CPs were lost to expiry by the end of the delivery window (27).In London’s polio campaign 24% of ordered doses were unused in CPs, compared to 14% in participating vaccination centres (24). The East of England noted higher RSV vaccine wastage in CPs (23% unused) than in GPs (15% unused), and the overall delivery volume was 3.5 times higher in GP than CP (26).

### Inequalities

Eight of the 14 included studies reported data on vaccine inequalities (2, 4, 15, 16, 19, 20, 23, 28). Research in Wales and England highlighted that females were more likely to receive the flu vaccination in CPs than men (20, 28). In addition, pharmacies perceived their service as important to access multiethnic urban populations by leveraging existing community networks (16). A report from the Company Chemists Association suggested that more than half of all COVID-19 vaccinations in the UK delivered to Black/Black British (55%), Asian/ Asian British (62%), mixed (54%) and Chinese or other ethnic groups (58%) in Spring 2024 were delivered in community pharmacies – although, as described above, the methodology and data sources underpinning these estimates are not clearly specified in the report and cannot be verified (23).

Research on whether CPs added value to the GP model, by offering broadening access to underserved populations was variable. Data from London boroughs showed that while 99% of CP flu vaccine recipients were registered with a GP, up to 24% of patients received their vaccine outside the geographical area covered by the body that commissioned health services in their local area at that time (2). Additionally, in Wales 24.9% of CP flu vaccination patients in 2013-14 had not been vaccinated the previous year (20), although it is unclear whether other factors may have contributed to this (e.g. changing eligibility criteria). However, data from the West Midlands showed a lower proportion of new CP patients receiving the flu vaccine (7.9%) compared to other research conducted in the UK, suggesting replacement rather than expansion of services (19). CP-based flu vaccination delivery in Wales covered a higher proportion of younger at-risk individuals than the overall flu programme. The proportion of at-risk individuals under 65 vaccinated in CPs (47.5%) was significantly higher than in GPs (28.6%) and carers accounted for nearly a quarter of CP-administered flu doses (20). In southwest England, childhood flu vaccine uptake declined with increasing socioeconomic deprivation, whether provided through school-based, GP or CP delivery programmes. CP and GP programmes also had significantly higher site-to-site variation after adjusting for deprivation, suggesting that school-based programmes were more effective at responding to the geographical and environmental factors beyond deprivation that may influence vaccine uptake (4).

## Discussion

### Main findings of the study

While use of CP delivery has become increasingly common for some vaccination programmes (principally seasonal programme such as influenza and COVID-19) over time, the number of vaccinations that are being delivered in CP settings across the UK remains small relative to that of GPs who provide the core service. There is currently no evidence to suggest CP delivery improves vaccination uptake. Research from the West Midlands and London suggests that the introduction of CPs as vaccination providers did not increase influenza vaccine coverage rates. The contrary finding from Wales which indicated an increase in the proportion of influenza vaccinations delivered by CPs, from 0.28% to 5.7%, without a corresponding decrease in GP activity, did not however result in increased coverage. These findings contrast with those from studies elsewhere, including the United States – likely explained by considerable differences in health system organisation and financing (32).

High levels of CP user satisfaction were consistently reported, with convenience, walk-in availability, and extended opening hours frequently cited as access enablers. The absence of comparative data on GP user satisfaction, however, makes it difficult to draw conclusions. These findings are consistent with previous literature which identified convenience and accessibility as central to patient acceptability of pharmacy-based vaccination services (32, 33). These factors are mirrored in findings on public acceptability of primary care services (34–37). Other research reported that most vaccinations given by CPs occurred during working hours (28) suggesting that longer opening hours do not translate into additional vaccination opportunities. Tension between CPs and GPs were noted across a number of studies, driven by concerns about vaccine data flows, patient safety, and financial implications.

This review highlighted the need for cost effectiveness data on CPs as a vaccine delivery model. Included studies universally reported costs rather than cost-effectiveness. Comparative costs incurred by CP and GP delivery models need to be recorded and should account for a pre-existing requirement for GPs to cover the costs of call-recall and vaccination booking activities. The grey literature emphasized logistical challenges faced by CPs, particularly concerns regarding high rates of vaccine wastage. Hence, a comprehensive cost effectiveness analysis that accounts for the level of support and logistics of the CP delivery model is required to assess whether the CP delivery model is effective, sustainable and scalable.

Sociodemographic inequalities are known to contribute significantly to variations in vaccination uptake (38–40), however our review did not find any evidence to suggest that CPs in the UK can reach populations that would not otherwise access vaccination via existing delivery routes. The only document to report uptake by ethnicity did not describe the methodology by which reported statistics were generated and industry sponsorship requires due caution of this analysis (23). There was also no evidence from studies reviewed to indicate that CPs increase uptake among groups known to have lower uptake for national programmes (e.g. those in clinical risk groups, and flu vaccination uptake among women who are pregnant). Our review found that CPs were perceived by some stakeholders to be well placed to reach populations that typically experience access barriers, because of their locations and links to community groups (3, 9). However, this is also the case for other primary care service delivery models and the lack of direct, comparative data between CP and GP vaccination delivery models precludes definitive assessment of their relative effectiveness to reach under-vaccinated populations. There is also good evidence that, for example, certain ethnic minority populations view GPs as the key point for credible information, particularly those who have limited English and digital literacy (37).

In the absence of comparative data on uptake, acceptability and cost-effectiveness for different delivery models, it remains challenging to determine how, where and for which populations CPs offer greatest value in supporting vaccination delivery. The balance of current evidence suggests they may have value in supporting time-limited activities including seasonal vaccination campaigns, but reported wastage rates and capacity constraints suggest against a role for CPs in lower-volume delivery (e.g. for routine immunisation programmes); and their impact in addressing immunisation inequities remains unproven.

### Strengths and limitations

This review has provided updated evidence on the potential value of using CPs as delivery models for delivering vaccinations across the UK, providing a critical assessment of evidence on their impact on national vaccinations which is essential to inform future policy and funding decisions. It also highlights that there is a need for further research and evaluation work on CPs as a vaccine delivery model prior to any proposed wider rollout.

Limitations of this evidence review include a focus exclusively on evidence from the UK, noting that insights from other settings – while informative – are often hampered by questions of transferability. A UK focus in itself elides significant differences in service delivery models and broader system contexts across the devolved nations which cannot for reasons of space be fully captured here, but which may have influenced outcomes reported in included studies. Many studies were very limited in scope, with data on either one pharmacy chain or only including pharmacies from a very specific locality. In addition, included peer-reviewed studies varied considerably in their methodological robustness and in terms of quality of reporting. Multiple papers had low survey response rates resulting in a high chance of response bias and inability to generalize findings to a broader population; papers also tended to report using different outcome measures reducing comparability of findings.

Grey literature sources also varied considerably in how they reported data. The tendency to report raw uptake figures without comparators from other relevant service routes (e.g. general practice) is a significant limitation on interpretation of findings. For example, 27 doses were administered to CP attendees aged 5-11 years via the northwest England MMR pilot reported above (27) – but this figure is difficult to interpret without the broader context that around 4,000 first dose MMR vaccinations administered to children aged 6-11 living in the catchment of the Greater Manchester Integrated Care Board during MMR catch-up activities in 2023-24 overall (41). There was also wide variation in depth of reporting: for some pilot programmes, such as Mpox and RSV, there were formal evaluation reports to draw findings from (albeit without clear reporting of methodological limitations) (25, 26), but for Polio, only supply data were provided with limited context on how the vaccination program was implemented or perceived by stakeholders (24), and data were sometimes reported at interim points rather than at completion of the relevant pilot so may not have reflected final findings.

In addition, whilst the published literature predominantly emphasized the acceptability of CP-based delivery by patients using, and pharmacists delivering, the services, grey literature sources were more likely to highlight procedural, logistical, and operational barriers encountered during the implementation of vaccination programmes in CPs. Despite the importance of understanding the logistical aspects of on-the-ground implementation, published literature offered limited insight into these areas. Specific details such as vaccine storage requirements, staffing considerations, patient invitation processes, and data reporting were often not included in published evaluations.

## Conclusion

This systematic review examined the contribution of CPs to vaccination programmes in the UK between 2015 and 2025. The available evidence shows that CPs do not increase overall vaccination uptake or effectively reach populations that would not otherwise access immunization through general practice. While CPs are generally viewed as acceptable and convenient by users, there is insufficient evidence to assess their cost-effectiveness, with existing studies failing to account for both direct and indirect costs. These findings suggest that while CPs may offer a convenient alternative for some patients, the evidence of their broader value in expanding routine vaccination coverage or improving equity remains limited.

## Acknowledgements

None.

## Competing Interests

There are no competing interests to declare.

## Data Availability

All included published literature in this systematic review is publicly available.

## Ethics

As this study was based on secondary analysis of published material, ethical approval was not required.

## Funding

This study is funded by the National Institute for Health and Care Research (NIHR) Health Protection Research Unit in Vaccines and Immunisation (NIHR200929), a partnership between UK Health Security Agency (UKHSA) and the London School of Hygiene and Tropical Medicine. The views expressed are those of the author(s) and not necessarily those of the NIHR, UKHSA or the Department of Health and Social Care.

## Patient and Public Involvement

As this study was a systematic review of existing literature, patients and the public were not directly involved in the design, conduct, or reporting of the research. The review did not involve primary data collection, recruitment, or the development of outcome measures. Findings may inform future research or service evaluations that include public and patient involvement, particularly in relation to the delivery and acceptability of vaccination services.

## Notes

### Competing Interest Statement

The authors have declared no competing interest.

### Summary of Updates

This version of the manuscript has been updated to include a more comprehensive search of existing papers. The previous manuscript only included a search of MEDLINE, this manuscript includes additional searches from Embase, Scopus, Web of Science, and CINAHL. Searching these additional databases provided us with two additional papers that are now included in this revised manuscript, the changes to the results were very minimal.

