## Supplemental File 1 for "Do community pharmacies add value to immunization programs? A systematic review from the UK between 2015-2025"

**Supplementary File 1: Database search strategies**

**MEDLINE SEARCH STRATEGY**

1	exp Great Britain/

2	(national health service* or nhs*).ti,ab,in.

3	(english not ((published or publication* or translat* or written or language* or speak* or literature or citation*) adj5 english)).ti,ab.

4	(gb or "g.b." or britain* or (british not "british columbia") or uk or "u.k." or united kingdom* or (england* not "new england") or northern ireland* or northern irish* or scotland* or scottish* or ((wales or "south wales") not "new south wales") or welsh*).ti,ab,jw,in.

5	(bath or birmingham or bradford or brighton or bristol or cambridge or canterbury or chester or chichester or coventry or derby or durham or exeter or gloucester or hereford or hull or lancaster or leeds or leicester or lincoln or liverpool or london or manchester or newcastle or norwich or nottingham or oxford or peterborough or plymouth or portsmouth or preston or salford or salisbury or sheffield or southampton or st albans or stoke or sunderland or truro or wakefield or wells or westminster or winchester or wolverhampton or worcester or york).ti,ab,in.

6	(bangor or cardiff or newport or st asaph or st davids or swansea).ti,ab,in.

7	(aberdeen or dundee or edinburgh or glasgow or inverness or perth or stirling).ti,ab,in.

8	(armagh or belfast or lisburn or londonderry or derry or newry).ti,ab,in.

9	1 or 2 or 3 or 4 or 5 or 6 or 7 or 8

10	(exp africa/ or exp americas/ or exp antarctic regions/ or exp arctic regions/ or exp asia/ or exp oceania/) not (exp great britain/ or europe/)

11	9 not 10

12	Community Pharmacy Services/

13	(pharmacist or pharmacy or pharmacies or chemist).ti,ab,kw

14	12 or 13

15	Vaccination/ or Immunization/ or Immunization programmes/

16	(Vacci* or Immuni*).ti,ab,kw

17	15 or 16

18	(Flu or Influenza or Covid-19 or SARS-Cov-2 or MMR or Measles or Mumps or Rubella or Poliovirus or Polio or OPV or IPV or RSV or "respiratory syncytial virus" or rotavirus or meningitis or meningococcal or MenACWY or MenB or MenC or pneumococcal or Hib or HPV or "Human papillomavirus" or Shingles or "Whooping Cough" or Pertussis or Routine or childhood).ti,ab,kw

19	11 and 14 and 17 and 18

20	19 and (2015 Nov* or 2015 Dec*).dp.

21	limit 19 to yr="2016 - Current"

22	20 or 21

**EMBASE SEARCH STRATEGY**

1 exp United Kingdom/

2 (national health service* or nhs*).ti,ab,in.

3 (english not ((published or publication* or translat* or written or language* or speak* or literature or citation*) adj5 english)).ti,ab.

4 (gb or "g.b." or britain* or (british not "british columbia") or uk or "u.k." or united kingdom* or (england* not "new england") or northern ireland* or northern irish* or scotland* or scottish* or ((wales or "south wales") not "new south wales") or welsh*).ti,ab,kw,in.

5 (bath or birmingham or bradford or brighton or bristol or cambridge or canterbury or chester or chichester or coventry or derby or durham or exeter or gloucester or hereford or hull or lancaster or leeds or leicester or lincoln or liverpool or london or manchester or newcastle or norwich or nottingham or oxford or peterborough or plymouth or portsmouth or preston or salford or salisbury or sheffield or southampton or st albans or stoke or sunderland or truro or wakefield or wells or westminster or winchester or wolverhampton or worcester or york).ti,ab,kw,in.

6 (bangor or cardiff or newport or st asaph or st davids or swansea).ti,ab,kw,in.

7 (aberdeen or dundee or edinburgh or glasgow or inverness or perth or stirling).ti,ab,kw,in.

8 (armagh or belfast or lisburn or londonderry or derry or newry).ti,ab,kw,in.

9 1 or 2 or 3 or 4 or 5 or 6 or 7 or 8

10 (exp africa/ or exp america/ or exp antarctica/ or exp arctic region/ or exp asia/ or exp oceania/) not (exp united kingdom/ or europe/)

11 9 not 10

12 community pharmacy/ or pharmacies/ or pharmacists/ or community pharmacist/

13 (pharmacist or pharmacy or pharmacies or chemist).ti,ab,kw.

14 12 or 13

15 vaccination/ or immunization/ or immunization program/

16 (vacci* or immuni*).ti,ab,kw.

17 15 or 16

18 (Flu or Influenza or Covid-19 or SARS-Cov-2 or MMR or Measles or Mumps or Rubella or Poliovirus or Polio or OPV or IPV or RSV or "respiratory syncytial virus" or rotavirus or meningitis or meningococcal or MenACWY or MenB or MenC or pneumococcal or Hib or HPV or "Human papillomavirus" or Shingles or "Whooping Cough" or Pertussis or Routine or childhood).ti,ab,kw.

19 11 and 14 and 17 and 18

20 19 and (2015 Nov* or 2015 Dec*).dp.

21 limit 19 to yr="2016 - Current"

22 20 or 21

**SCOPUS SEARCH STRATEGY**

TITLE-ABS-KEY ( ( "national health service" OR nhs OR gb OR "g.b." OR britain* OR british OR uk OR "u.k." OR "united kingdom" OR england OR scotland OR wales OR "northern ireland" OR bath OR birmingham OR bradford OR brighton OR bristol OR cambridge OR canterbury OR chester OR chichester OR coventry OR derby OR durham OR exeter OR gloucester OR hereford OR hull OR lancaster OR leeds OR leicester OR lincoln OR liverpool OR london OR manchester OR newcastle OR norwich OR nottingham OR oxford OR peterborough OR plymouth OR portsmouth OR preston OR salford OR salisbury OR sheffield OR southampton OR "st albans" OR stoke OR sunderland OR truro OR wakefield OR wells OR westminster OR winchester OR wolverhampton OR worcester OR york OR bangor OR cardiff OR newport OR "st asaph" OR "st davids" OR swansea OR aberdeen OR dundee OR edinburgh OR glasgow OR inverness OR perth OR stirling OR armagh OR belfast OR lisburn OR londonderry OR derry OR newry ) AND ( pharmacist* OR pharmacy OR pharmacies OR chemist OR chemists ) AND ( vacci* OR immuni* OR "immunization program*" OR "immunisation program*" OR vaccination OR immunization ) AND ( flu OR "covid-19" OR "sars-cov-2" OR mmr OR measles OR mumps OR poliovirus OR polio OR opv OR ipv OR rsv OR "respiratory syncytial virus" OR rotavirus OR meningitis OR meningococcal OR menacwy OR menb OR menc OR pneumococcal OR hib OR hpv OR "human papillomavirus" OR shingles OR "whooping cough" OR pertussis OR routine OR childhood ) ) AND NOT TITLE-ABS-KEY ( "new england" OR "new york" OR "british columbia" OR "new south wales" OR ontario OR toronto OR massachusetts OR boston OR harvard OR nebraska OR australia OR zealand OR nsw ) AND PUBYEAR > 2014 AND PUBYEAR < 2026

**WEB OF SCIENCE SEARCH STRATEGY**

TS=
((“national health service” OR nhs OR gb OR "g.b." OR britain* OR british OR uk OR "u.k." OR "united kingdom" OR england OR scotland OR wales OR "northern ireland" OR bath OR birmingham OR bradford OR brighton OR bristol OR cambridge OR canterbury OR chester OR chichester OR coventry OR derby OR durham OR exeter OR gloucester OR hereford OR hull OR lancaster OR leeds OR leicester OR lincoln OR liverpool OR london OR manchester OR newcastle OR norwich OR nottingham OR oxford OR peterborough OR plymouth OR portsmouth OR preston OR salford OR salisbury OR sheffield OR southampton OR "st albans" OR stoke OR sunderland OR truro OR wakefield OR wells OR westminster OR winchester OR wolverhampton OR worcester OR york OR bangor OR cardiff OR newport OR "st asaph" OR "st davids" OR swansea OR aberdeen OR dundee OR edinburgh OR glasgow OR inverness OR perth OR stirling OR armagh OR belfast OR lisburn OR londonderry OR derry OR newry)

AND

(pharmacist* OR pharmacy OR pharmacies OR chemist OR chemists)

AND

(vacci* OR immuni* OR "immunization program*" OR "immunisation program*" OR vaccination OR immunization)

AND

(flu OR influenza OR covid-19 OR sars-cov-2 OR mmr OR measles OR mumps OR rubella OR poliovirus OR polio OR opv OR ipv OR rsv OR "respiratory syncytial virus" OR rotavirus OR meningitis OR meningococcal OR menacwy OR menb OR menc OR pneumococcal OR hib OR hpv OR "human papillomavirus" OR shingles OR "whooping cough" OR pertussis OR routine OR childhood))

NOT TS=("new england" OR “new york” OR "british columbia" OR "new south wales" OR ontario OR toronto OR massachusetts OR boston OR harvard OR nebraska OR australia OR zealand OR nsw)

Date Set: 2015 - current

**CINAHL SEARCH STRATEGY**

S1 (MH "Great Britain") OR (MH "United Kingdom")

S2 TI (national health service* OR nhs*) OR AB (national health service* OR nhs*)

S3 TI (english NOT ((published OR publication* OR translat* OR written OR language* OR speak* OR literature OR citation*) N5 english)) OR AB (english NOT ((published OR publication* OR translat* OR written OR language* OR speak* OR literature OR citation*) N5 english))

S4 TI (gb OR "g.b." OR britain* OR (british NOT "british columbia") OR uk OR "u.k." OR "united kingdom" OR (england* NOT "new england") OR "northern ireland" OR "northern irish" OR scotland* OR scottish* OR ((wales OR "south wales") NOT "new south wales") OR welsh*)

S5 TI (bath OR birmingham OR bradford OR brighton OR bristol OR cambridge OR canterbury OR chester OR chichester OR coventry OR derby OR durham OR exeter OR gloucester OR hereford OR hull OR lancaster OR leeds OR leicester OR lincoln OR liverpool OR london OR manchester OR newcastle OR norwich OR nottingham OR oxford OR peterborough OR plymouth OR portsmouth OR preston OR salford OR salisbury OR sheffield OR southampton OR "st albans" OR stoke OR sunderland OR truro OR wakefield OR wells OR westminster OR winchester OR wolverhampton OR worcester OR york) OR AB (bath OR birmingham OR bradford OR brighton OR bristol OR cambridge OR canterbury OR chester OR chichester OR coventry OR derby OR durham OR exeter OR gloucester OR hereford OR hull OR lancaster OR leeds OR leicester OR lincoln OR liverpool OR london OR manchester OR newcastle OR norwich OR nottingham OR oxford OR peterborough OR plymouth OR portsmouth OR preston OR salford OR salisbury OR sheffield OR southampton OR "st albans" OR stoke OR sunderland OR truro OR wakefield OR wells OR westminster OR winchester OR wolverhampton OR worcester OR york)

S6 TI (bangor OR cardiff OR newport OR "st asaph" OR "st davids" OR swansea) OR AB (bangor OR cardiff OR newport OR "st asaph" OR "st davids" OR swansea)

S7 TI (aberdeen OR dundee OR edinburgh OR glasgow OR inverness OR perth OR stirling) OR AB (aberdeen OR dundee OR edinburgh OR glasgow OR inverness OR perth OR stirling)

S8 TI (armagh OR belfast OR lisburn OR londonderry OR derry OR newry) OR AB (armagh OR belfast OR lisburn OR londonderry OR derry OR newry)

S9 S1 OR S2 OR S3 OR S4 OR S5 OR S6 OR S7 OR S8

S10 (MH "Africa+") OR (MH "Americas+") OR (MH "Antarctic Regions+") OR (MH "Arctic Regions+") OR (MH "Asia+") OR (MH "Oceania+") NOT ( (MH "Great Britain+") OR (MH "Europe+") )

S11 S9 NOT S10

S12 (MH "Community Pharmacies+") OR (MH "Pharmacists+")

S13 TI (pharmacist OR pharmacy OR pharmacies OR chemist) OR AB (pharmacist OR pharmacy OR pharmacies OR chemist)

S14 S12 OR S13

S15 (MH "Immunization+") OR (MH "Vaccination+") OR (MH "Immunization Programs+")

S16 TI (vacci* OR immuni*) OR AB (vacci* OR immuni*)

S17 S15 OR S16

S18 TI (flu OR influenza OR covid-19 OR sars-cov-2 OR mmr OR measles OR mumps OR rubella OR poliovirus OR polio OR opv OR ipv OR rsv OR "respiratory syncytial virus" OR rotavirus OR meningitis OR meningococcal OR menacwy OR menb OR menc OR pneumococcal OR hib OR hpv OR "human papillomavirus" OR shingles OR "whooping cough" OR pertussis OR routine OR childhood) OR AB (flu OR influenza OR covid-19 OR sars-cov-2 OR mmr OR measles OR mumps OR rubella OR poliovirus OR polio OR opv OR ipv OR rsv OR "respiratory syncytial virus" OR rotavirus OR meningitis OR meningococcal OR menacwy OR menb OR menc OR pneumococcal OR hib OR hpv OR "human papillomavirus" OR shingles OR "whooping cough" OR pertussis OR routine OR childhood)

S19 S11 AND S14 AND S17 AND S18
